# Long-Term Remodeling Response in the Lamina Cribrosa Years after IOP Lowering by Suturelysis after Trabeculectomy

**DOI:** 10.1101/2023.05.30.23290730

**Authors:** Cameron A. Czerpak, Harry A. Quigley, Thao D. Nguyen

**Author notes:** **Corresponding Author:** Cameron A. Czerpak, Department of Mechanical Engineering, Whiting School of Engineering, Johns Hopkins University, 3400 N. Charles St., 125 Latrobe Hall, Baltimore, Maryland USA 21218. **Financial Support:** This research was funded in part by NIH NEI R01 02120 (Dr. Quigley), Brightfocus G2021012S, P30 01765 (Wilmer Institute Core Grant), Research to Prevent Blindness, the A. Edward Maumenee Professorship (Dr. Quigley), and BrightFocus. **Supplemental Information:** This article contains additional online-only material. The following should appear online-only: Supplement Tables S1-S21.

## Abstract

**Objective:** To measure the active remodeling of the lamina cribrosa (LC) years after IOP lowering by suturelysis.

**Design:** Cohort study.

**Participants:** Glaucoma patients were imaged 20 minutes after laser suturelysis following trabeculectomy surgery and at their follow-up appointment 1-4 years later (16 image pairs; 15 persons).

**Intervention:** Non-invasive optical coherence tomography (OCT) imaging of the eye.

**Main Outcomes:** Deformation calculated by correlating OCT scans of the LC immediately after IOP lowering by suturelysis and those acquired years later (defined as *follow-up* strain).

**Results:** Mean LC follow-up strain in the anteroposterior direction (*E_zz_*) was 14.0 ± 21.3% (mean, standard deviation, p=0.03), while the LC anterior border moved 60.9 ± 54.6 μm into the eye (p=0.0006) on long-term, maintained IOP lowering. *E_zz_* at follow-up was 14 times larger than the direct *E_zz_* response to IOP lowering by suturelysis. There was a significant association between larger LC anterior movement and greater *E_zz_* (p=0.004) at follow-up. Thinner retinal nerve fiber layer (RNFL) at suturelysis was associated with greater follow-up *E_zz_* (p=0.04). Worsening visual field indexes during follow-up were associated with greater LC widening (positive remodeling *E_θθ_*, p=0.02). Eyes with a greater counterclockwise twist (positive *E_θz_*) at suturelysis had greater reversal clockwise twist at follow-up (negative remodeling *E_θz_*, p=0.007).

**Conclusion:** Follow-up strains and LC border position changes measured years after IOP lowering are far larger than the immediate strain response and LC border movement response to IOP lowering and indicate dramatic remodeling of the LC anatomical structure caused by IOP lowering and glaucoma progression. The remodeling includes a substantial increase in LC thickness and movement into the eye. Eyes with greater direct strain response to IOP-lowering strains, greater glaucoma damage at suturelysis, and greater worsening of visual field at follow-up experienced greater remodeling

**Trial Registration:** ClinicalTrials.gov Identifier: NCT03267849

## INTRODUCTION

The lamina cribrosa (LC) is the primary site of retinal ganglion cell axon damage in glaucoma within the optic nerve head (ONH).^1–4^ Increases in intraocular pressure (IOP) from glaucoma increase biomechanical stress at the ONH, producing deformations that can be quantified by measuring strains, which estimate expansions, contractions, and twisting motions. Strains in the ONH can lead to detrimental effects on astrocytes, axons, and nutritional blood flow.^5–9^ The ONH response to IOP may provide biomarkers for the incidence and progressive worsening of glaucoma.^10^

The structures and collagen network in the ONH have been shown to remodel in human glaucoma eyes^11–13^ and experimental monkey glaucoma.^14–17^ The LC alterations include a thinner, more curved, and more excavated shape.^18^ The pores within the LC network are filled in by astrocytes that migrate into pores.^19^ IOP affects the cells within the ONH, and its neurons, glia and fibroblasts have been shown to have mechanosensitive properties.^20–24^

Glaucoma treatment to reduce IOP has been demonstrated to be beneficial, but the cellular events that occur in the ONH after IOP lowering have not been studied in detail. We and others have conducted *in vivo* research to study the events following IOP lowering using optical coherence tomography (OCT). Sequential OCT imaging can measure ONH mechanical behavior by comparing images at higher and lower IOP. Many previous studies measured only the depth change of the LC anterior border^25^ after IOP change, demonstrating that LC depth decreases weeks to months after IOP lowering.^26–28^ Shortly after an average of 12 mmHg of IOP lowering, we found that the LC moved an average of 1.33 ± 6.26 µm posteriorly (out of the eye) when measured 20 minutes after suturelysis following trabeculectomy surgery. The time sequence of LC border movement after trabeculectomy, studied by Kadziauskiene et al.^29^, was greatest 3-10 days after trabeculectomy (40 μm anterior), followed by a slower, further anterior LC border movement during the next 6 months, as also found by Krzyżanowska-Berkowska et al.^30, 31^ and others.^32^ This suggests that the LC border experiences at least 2 phases of change after IOP lowering: an initial period of passive biomechanics, potentially due to viscoelastic response of its extracellular matrix, followed by a longer, slower period of tissue remodeling involving cellular responses.

Thus, the movement of the LC border is a limited surrogate for the dynamic events affecting LC mechanics. Deformations within the LC tissues may provide more accurate assessment of stretching, twisting, and compression that retinal ganglion cell axons experience. We and others have quantified the mechanical deformations from short-term IOP change within the optic nerve by measuring strain using digital volume correlation (DVC).^33–42^ In this time frame, the LC expanded in thickness by 1% and contracted in radius by 0.2%. By contrast, Girard et al.^37^ measured LC strains of 8.6% at 50 days after IOP lowering. No study to date measured mechanical ONH strains years after IOP lowering. In this study, we utilized DVC analysis to compare ONH images taken immediately after IOP lowering by suturelysis with images of the same eyes 1-4 years later, producing what are defined here as remodeling strains of the internal LC tissue.

## METHODS

### Experimental Participants

This study was approved by the institutional review board of the Johns Hopkins School of Medicine and can be found at ClinicalTrials.gov as NCT03267849. Written informed consent was obtained from every patient prior to OCT imaging and IOP measurements and the study adheres to the tenets of the Declaration of Helsinki.

Sixteen eyes from 15 patients were reimaged 1-4 years (2.04 ± 0.98 years) after IOP lowering using spectral-domain OCT (SD-OCT, Spectralis, Heidelberg Engineering, Heidelberg, Germany) and the same procedure as used in the prior published study.^42^ All patients had undergone trabeculectomy and a postoperative suturelysis procedure and participated in our previous study.^42^ Patient age at the time of suturelysis ranged from 54 to 86 years old (mean 67.6 ± 8.3 years). One patient underwent suturelysis in both the left and right eyes and both were included here. Mean deviation (MD) and visual field index (VFI) were captured at routine visits using Humphrey Field Analyzer (HFA) 24-2 Swedish Interactive Threshold Algorithm (SITA Standard) field tests (Zeiss Meditec, Dublin, CA) within 3 months of suturelysis imaging and the time of follow-up imaging. Retinal nerve fiber layer (RNFL) thickness was measured using Cirrus OCT (Zeiss) at the time of suturelysis. Fewer than half of the patients completed RNFL measurements at the time of the follow-up imaging and therefore change in RNFL over time was not included in the analysis. MD at suturelysis ranged from severe to mild damage (−27.43 to - 1.32 dB; mean (standard deviation) = −10.24 ± 6.82 dB). The distribution of damage was: 4 mild damage eyes (MD > −6), 6 moderate damage eyes (−12 dB < MD < −6 dB), and 6 severe damage eyes (MD < −12 dB). Mean visual field MD change over time from surgery to later imaging was −1.11 ± 3.98 dB.

### OCT Imaging

IOP was measured before each imaging session using the iCare tonometer (iCare Finland Oy). IOP after suturelysis ranged from 3 to 23 mmHg (12.88 ± 6.39 mmHg) and IOP at the 1-4 year follow up ranged from 3 to 19 mmHg (8.75 ± 4.58 mmHg), for a modest average decrease in IOP of 4.13 ± 7.89 mmHg (p=0.05). We also recorded applanation IOP measurements from up to 3 visits before trabeculectomy, with mean = 19.85 ± 7.07 mmHg, range: 7.33 – 37.0 mmHg. In addition, we recorded all IOP measurements from suturelysis to follow-up imaging (minimum, maximum, mean, range, standard deviation, and variance). Corneal curvature was measured using the IOL Master instrument (Zeiss Meditec) and input to Spectralis for each imaging session. Each patient was imaged using the ONH-RC scan pattern consisting of 24 radial SD-OCT scans with enhanced depth imaging on (Figure 1). The scans were centered on the ONH and were 768 x 495 pixels with 7.5° between adjacent scans in the radial (R), anterior-posterior (Z), and theta (Θ) directions, respectively. The resolution in R varied due to keratometry corrections (5.33-6.51 μm/pixel) and Z resolution remained constant at 3.87 μm/pixel. The resolution in 0 was approximately 108 μm/pixel at the BMO, with resolution improving towards the center of the LC. Three imaging sets were taken at the same session to estimate baseline and correlation errors using a previously published method.^34, 42^ Before processing with DVC, the natural speckle pattern of the OCT images was enhanced using contrast-limited adaptive histogram equalization (CLAHE) in FIJI (ImageJ) (Figure 1). Noise was reduced in areas of low signal using a gamma correction of 1.75 following CLAHE. Images were then imported into MATLAB 2019a (Mathworks) to reconstruct the images into a 3- dimensional matrix of 8-bit intensity values.

**Figure 1:**
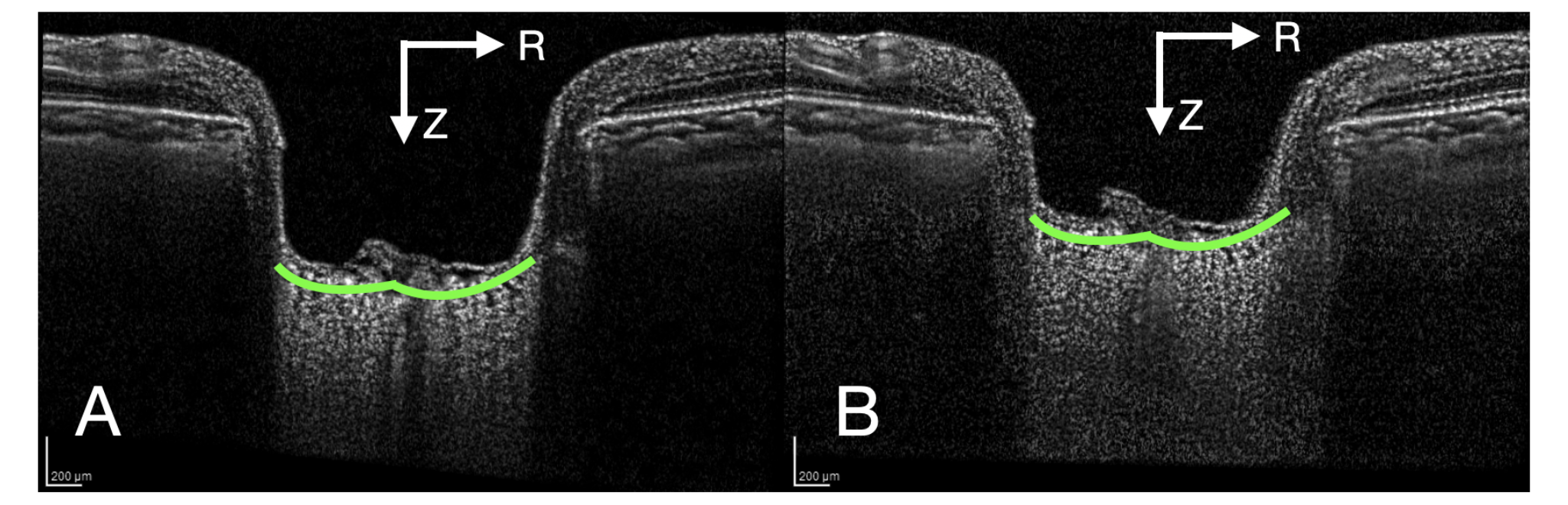
A) OCT image of the ONH after suturelysis, B) 1 year later.

### Image Segmentation

All 24 radial scans were manually segmented by a trained observer (C.A.C.) to mark the anterior border of the LC and Bruch’s membrane opening (BMO) on both sides of the ONH. The anterior LC markings were fit using piecewise linear interpolation, and a second parallel curvilinear border was drawn 250 μm posterior to represent the posterior LC. The LC region was defined as the area between the two LC borders and within BMO delineated by vertical lines.^42^ 250 μm was chosen as the posterior border because few eyes have a visible change in LC signal to clearly mark the posterior border of the LC. The chosen thickness was based on histological measurements of the human LC.^43^

### Digital Volume Correlation

We have published in detail the DVC method used to measure strain and strain error in the LC.^34, 42^ DVC was applied to correlate the OCT volumes acquired at the follow-up appointment years after suturelysis and the OCT volumes acquired 20 minutes after suturelysis to calculate the follow-up deformation. In contrast, the direct deformation response to IOP lowering was calculated by correlating the OCT volumes taken 20 minutes after suturelysis and the OCT volume acquired before suturelysis. The deformation at follow-up includes anatomical changes caused by the active remodeling response to glaucoma and the initial IOP lowering and by the direct deformation response to a mild decrease in IOP during the follow-up period.

DVC is a method that tracks the natural speckle pattern of the OCT images in three dimensions (R, Z, Θ) from a before image to an after image to measure mechanical deformations (displacements). The 3-D volume of the first image is broken down into subvolumes, and the algorithm searches the second volume for the best match to the before volume. At every location the algorithm tries to match the volumes, and it measures how well the volumes correlated (correlation-coefficient). The algorithm then records what displacements in R, Z, and Θ were needed to move the subvolume from the before to the after location with the highest correlation-coefficient. The DVC algorithm used in this study is an extension of Bar-Kochba et al.^44^ to analyze radial scans and calculate displacements *U_r_* (radial), *U_z_* (axial), and *U_θ_* (circumferential) in the cylindrical coordinate system. The algorithm subvolume begins large at 256 x 256 x 32 pixels (R, Z, Θ) and refines to 2 x 2 x 1 pixels to better measure large differences in displacements. A correlation coefficient indicates the quality of the subvolume matching, with low values primarily occurring due to image noise, low signal, or blood vessel shadows. A correlation coefficient of 0.055 was used to remove displacements from dark regions with no contrast.^34, 42^ The average percent of correlation in the LC was 49.2 ± 16% across the 16 image sets.

The output of DVC is a field of displacements. We then took the gradient of the displacement field to measure the relative deformations (expansions, contractions, and shears) within the field (strain). LC depth change following suturelysis was measured from the displacement of the anterior LC border *U_z_* with respect to the 2 BMO points. Displacements were averaged across the entire LC border, rather than choosing the central most point in the LC to measure displacement. Strain and LC depth change outcomes will be referred to as *follow-up*, meaning they measure the remodeling strains or displacements from after suturelysis to the *follow-up* imaging session years later. Strain and LC-depth change measured in our previous study^42^ with these eyes will be referred to as *after-lysis*, meaning they measure the strain from before suturelysis to 20 minutes *after suturelysis*.

The three normal (*E_zz_*, *E_rr_*, *E_θθ_*) and three shear strains (*E_zθ_*, *E_rz_*, *E_rθ_*) of the Green-Lagrange strain tensor were measured using DVC in 3-dimensions: anterior-posterior strain (*E_zz_*), radial strain (*E_rr_*), circumferential strain (*E_θθ_*), and shear strains cylindrical twist strain (*E_zθ_*), in plane shear strain (*E_rz_*), and shear strain that transforms a circular cross section into a twisted ellipse (*E_rθ_*). Positive values for normal strains represent elongation, while negative values represent contraction. Shear strains are deformations in two directions. A shear strain applied to a rectangle transforms the shape into a diamond. The maximum principal strain (*E_max_*) and maximum shear strain (*Γ_max_*) were calculated in the RZ plane (within the radial scan) to reduce the impact of the lower resolution of image data between the radial slices.

The potential error in strains were estimate in two ways: baseline error and correlation error. Baseline error is calculated by comparing 2 image sets taken immediately after the other and applying DVC. Ideally, baseline error would be 0; however, image quality and patient movement or misalignment produce some disparity. Correlation error was measured by manually applying a known displacement to an OCT image, then strains are calculated between the original and distorted images, and DVC applied. The difference between strain calculated from the known distortion and that after applying the displacement is the correlation error, again ideally 0. However, low image quality and the presence of blood vessels overlying the LC contribute to correlation error.

For the present images, the average overall baseline and correlation errors (*U_z_*) were −0.21 ± 0.73 μm and 0.62 ± 0.12 μm, respectively (Supplement Tables S1, S2). The average baseline and correlation strain errors for strain *E_zz_* were 0.134 ± 0.167% and 0.531 ± 0.122%, and for *E_rr_* were 0.021 ± 0.106% and −0.118 ± 0.079%, respectively. The strains and error equations have previously been published.^34, 42^

One patient had sustained low IOP (mean = 4 mmHg) with stable function despite some chorioretinal wrinkling, which required use of radial scans from two separate follow-up imaging sets to register with the after-lysis scans. Measurement of LC border movement was excluded for this patient as the measurement relies on the BMO position.

### Analysis

We used t-tests to determine the significance of the mean LC follow-up strains compared to no change and to baseline and correlation errors. Relationships between the follow-up strain, IOP, LC border movement, glaucoma damage (MD, VFI, and RNFL thickness), and after-lysis strain were analyzed using linear regression.

## RESULTS

### LC strains and depth change

At follow-up, the LC border moved 60.9 +/- 54.6 μm into the eye (anterior) (p=0.0007) and LC follow-up *E_zz_* strain was 14.0 +/- 21% (tensile, or increased in thickness, p=0.02; Figure 1, Table 1). Follow-up *E_max_* and *ρ_max_* were predominately influenced by the large follow-up *E_zz_* measurement and were 17.7 +/- 20.3% (p=0.003) and 10.7 +/- 9.74% (p=0.0005), respectively. Only follow-up *E_zz_* outcomes were greater than baseline error (p=0.02) and correlation error (p=0.02) (Supplement Table S3). The other follow-up components of the strain tensor were 1 to 2 orders of magnitude smaller than follow-up *E_zz_* (Table 1). Follow-up radial strain *E_rr_* was nearly 0 (p=0.98) and the LC contracted circumferentially but not significantly (follow-up *E_θθ_* = −0.278 +/- 2.44%; p=0.66). Comparing the follow-up strains to the after-lysis strains published previously for this subset of eyes^42^ showed that the follow-up *E_zz_* was 14 times larger than after-lysis *E_zz_* (0=0.02) and the follow-up LC border anterior movement was 124 times larger than 20 minutes after-lysis (p=0.0007). Follow-up *E_max_* and *Γ_max_* were also 8 (p=0.003) and 6 (p=0.0005) times larger than after-lysis *E_max_* and *Γ_max_*.

**Figure 2:**
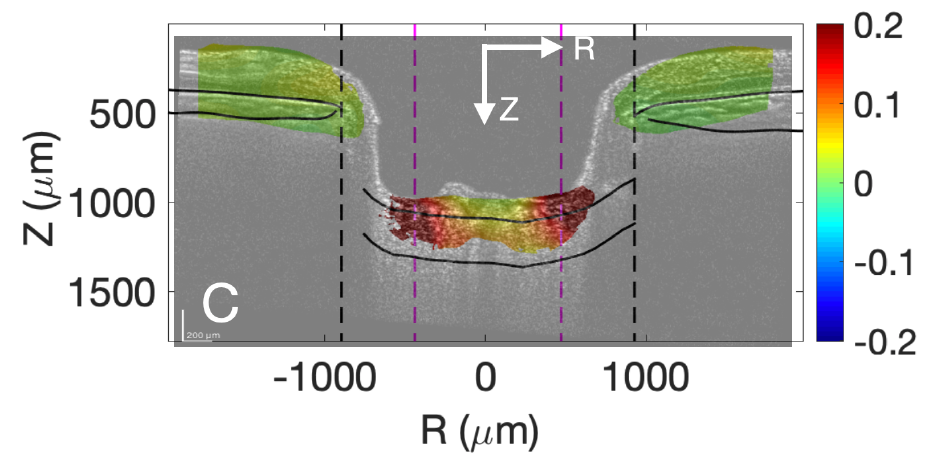
A representative nasal-temporal single B-scan from one patient illustrates a strongly positive follow-up *E_zz_*, indicating a 14% increase in LC thickness one year after IOP-lowering.

**Table 1:** Comparison of mean strains measured 20 minutes after suturelysis and follow-up strains from 1- 4 years later. ALD = anterior LC depth change (minus indicates anterior movement) (n=16 eyes).

|  | After-lysis<br>Mean | After-lysis vs<br>Follow-up<br>p-value | Follow-up<br>Mean | Follow-up<br>stdev | Follow-up > 0<br>p-value |
| --- | --- | --- | --- | --- | --- |
| *IOP (mmHg)* | 13 | **0.05** | 9 | 5 | — |
| *ALD ($\mu\text{m}$)* | 0.493 | **0.0006** | -60.9 | 54.6 | **0.0007** |
| $E_{zz}$ | 0.0104 | **0.03** | 0.140 | 0.213 | **0.02** |
| $E_{rr}$ | -0.00184 | 0.60 | 0.0000990 | 0.0138 | 0.98 |
| $E_{\theta\theta}$ | -0.00157 | 0.85 | -0.00278 | 0.0244 | 0.66 |
| $E_{r\theta}$ | 0.00191 | 0.43 | 0.00478 | 0.0129 | 0.16 |
| $E_{z\theta}$ | -0.000759 | 0.30 | -0.00613 | 0.0156 | 0.14 |
| $E_{rz}$ | 0.000110 | 0.47 | 0.00503 | 0.0261 | 0.45 |
| $E_{max}$ | 0.0214 | **0.009** | 0.177 | 0.203 | **0.003** |
| $\Gamma_{max}$ | 0.0171 | **0.003** | 0.107 | 0.0974 | **0.0005** |

### LC depth change with strain

We compared follow-up strains and follow-up LC border movement, which showed that the LC expands as the LC border moves into the eye (anterior). Greater movement of the LC border into the eye at follow-up was associated with more tensile follow-up *E_zz_* (R^2^=0.48, p=0.004), greater *E_max_* (R^2^=0.51, p=0.003), and greater *Γ_max_* (R^2^=0.54, p=0.002) (Figure 3) (Supplement Table S4). Follow-up LC border movement was not associated with follow-up *E_rr_*, *E_θθ_*, or shear strains (all p>0.11).

**Figure 3:**
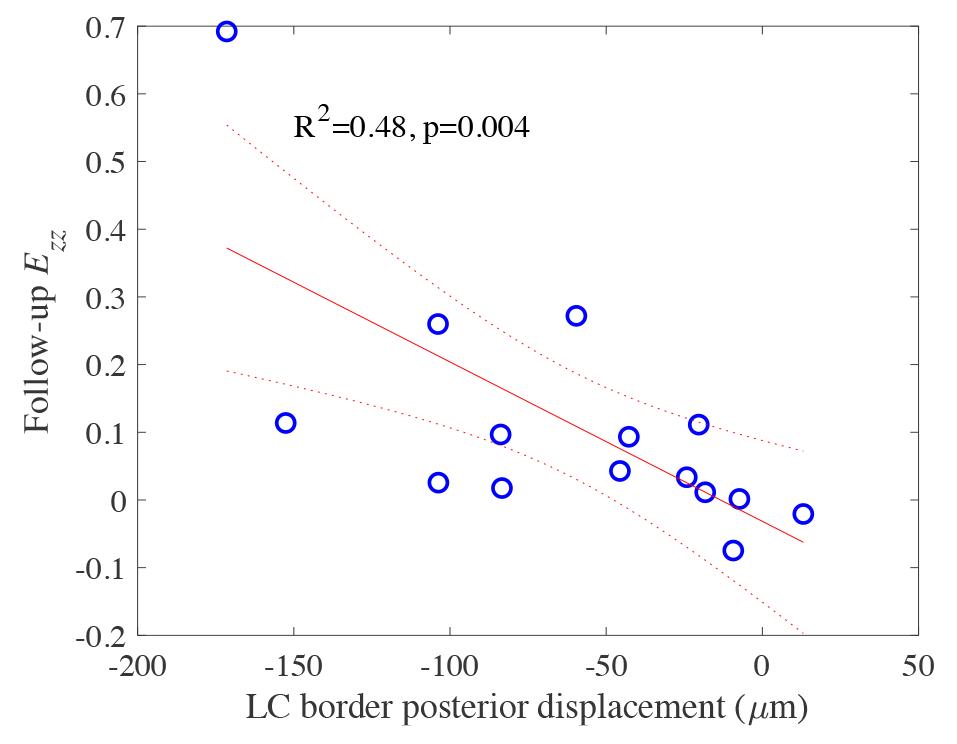
Linear regression analysis showed that greater anterior movement of the LC border was associated with greater follow-up *E_zz_* (LC thickening)

### LC after-lysis and IOP change

A smaller IOP decrease immediately after suturelysis was associated with greater follow-up movement of the LC border in the anterior direction (R^2^=0.27, p=0.046) (Supplement Table S5). We next compared the mean IOP for visits prior to trabeculectomy to the immediately after-lysis IOP and related this difference to LC outcomes. The IOP decrease from before trabeculectomy to after-lysis was not significantly associated with follow-up *E_θθ_* (R^2^=0.23, p=0.06), *E_rr_* (R^2^=0.20, p=0.08), or *E_zz_* (R^2^=0.0095, p=0.72) (Supplement Table S6). The IOP change from suturelysis to long-term follow-up was not associated with follow-up strain or LC border movement (p≥0.1) (Supplement Table S7). The time from suturelysis to follow-up imaging (1-4 years) was also not associated with the follow-up strain response (p>0.06) (Supplement Table S8). Regarding the IOP history at all appointments following suturelysis, neither maximum, minimum, range, mean, standard deviation nor variance of IOP were associated with follow-up strain or LC border movement (all p>0.06) (Supplement Tables S9- S14).

### LC after-lysis and glaucomatous neuropathy

We investigated the effects of glaucoma damage on the follow-up strain using the RNFL thickness, MD, and VFI at suturelysis, and the MD and VFI change from after-lysis to long-term follow-up. Eyes with more glaucoma damage (thinner RNFL) experienced greater follow-up *E_zz_* (more expansion; R^2^=0.24, p=0.05; Figure 4), while thinner RNFL was nearly significantly associated with more anterior movement of the LC border (R^2^=0.24, p=0.06) (Supplement Table S15). MD and VFI at suturelysis were not associated with follow-up strain or LC border movement (p>0.3) (Supplement Tables S16-S17). Patients with greater MD decrease from suturelysis to follow-up experienced more tensile follow-up *E_θθ_* (R^2^=0.31, p=0.02) and positive *E_rθ_* (R^2^=0.28, p=0.03) (Supplement Table S18). Patients with greater decrease in VFI from suturelysis to follow up experienced more tensile *E_θθ_* (R^2^=0.33, p=0.02) (Figure 5) (Supplement Table S19).

**Figure 4:**
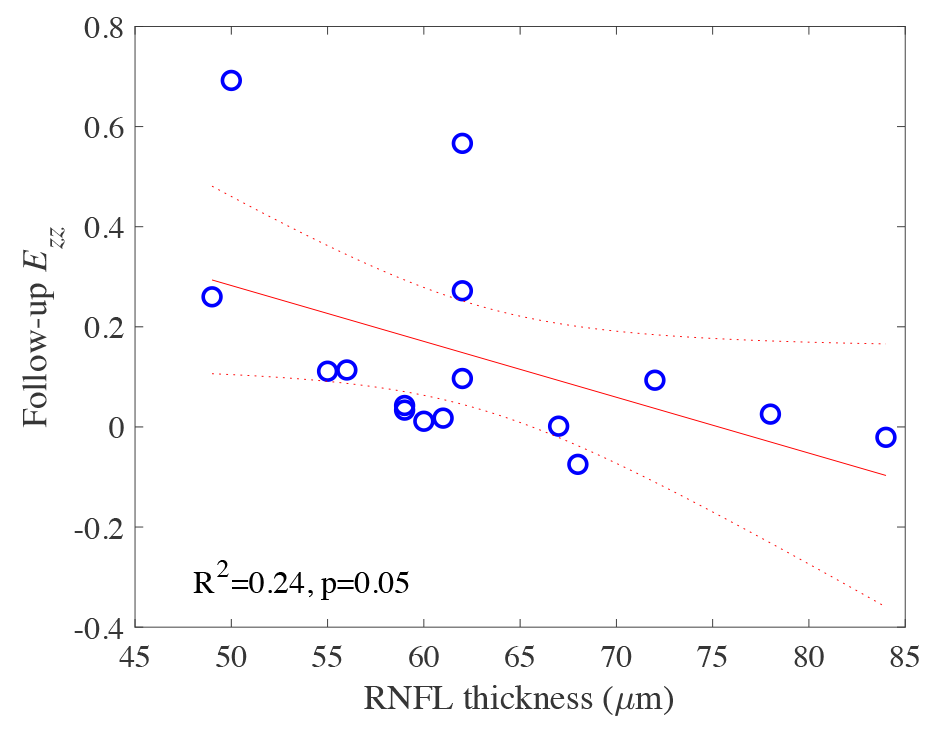
Eyes with more glaucoma damage (thinner RNFL) at suturelysis experienced greater follow-up *E_zz_* (LC thickening)

**Figure 5:**
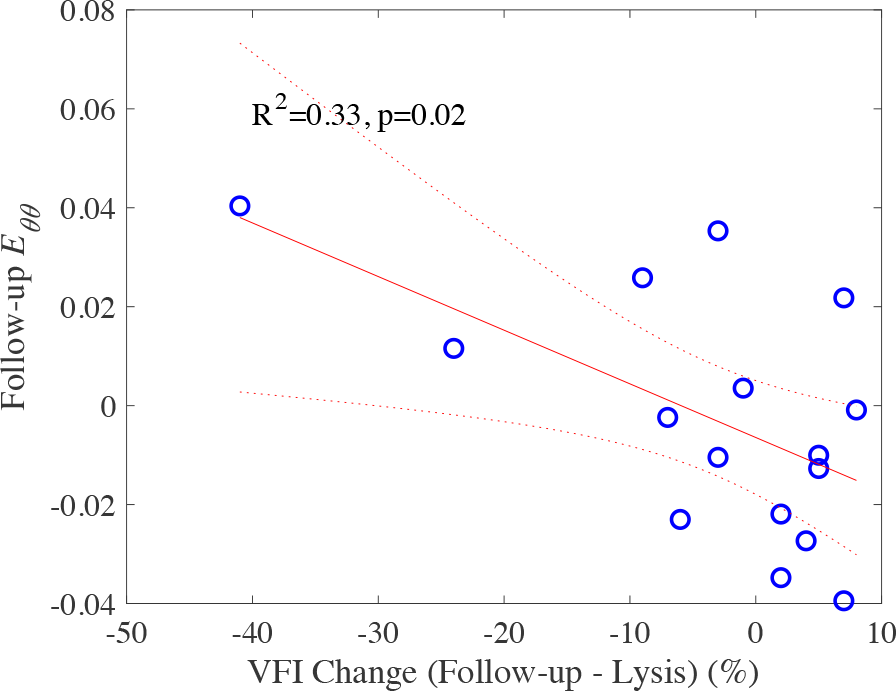
Patients with worse progression (decrease in VFI from suturelysis to follow-up) experienced more tensile follow-up *E_θθ_* (widening).

### Comparison of after-lysis and follow-up strains

Some after-lysis strains were associated with follow-up strains. Eyes with a greater counterclockwise twist (positive *E_θz_*) after-lysis had greater clockwise twist (negative *E_θz_*) at follow-up (R^2^=0.42, p=0.007) (Figure 6) (Supplement Table S20). Eyes with a greater after-lysis shear strain (*E_rθ_*) had a borderline association with greater MD decrease during the follow-up period (R^2^=0.16, p=0.12) (Supplement Table S21).

**Figure 6:**
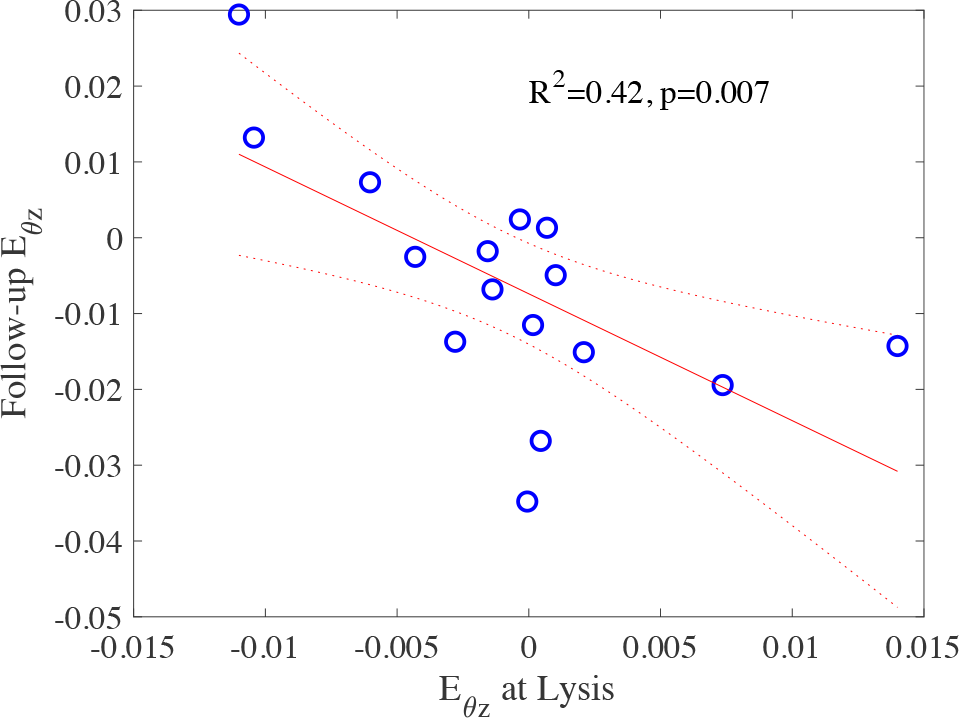
Eyes with a greater twist *E_θz_* had greater negative follow-up *E_θz_*.

## DISCUSSION

While many publications documented the dynamic nature of LC border movement after IOP lowering, only one study, to our knowledge, has measured LC strains at some time after IOP lowering.^37^ We measured strains both immediately after lowering and years later, with the hypothesis that detailed measurement of LC behavior throughout its structure would be more informative than documentation of its border alone, as has been frequently reported.^26, 45^ Our data show that follow-up LC strains were up to 14 times larger than after-lysis strains, with the most obvious increase in strain direction being anterior-posterior (*E_zz_*), indicating axial expansion. While follow-up radial, circumferential, and shear strains were larger than their after-lysis counterparts, they remained small compared to follow-up *E_zz_* strain.

We found that more movement of the LC border in the anterior direction was associated with larger *E_max_* and *Γ_max_*, similar to *E_zz_*. It is certainly consistent with intuitive logic that the LC would move anteriorly if it became thicker. However, as we previously reported^34, 42^, LC border movement is not associated with all components of strain. Furthermore, it is more likely that data on internal LC mechanical change would provide information on the expansions, contractions, and twisting motions transmitted to the axons that are thought to underlie glaucoma injury.

Strains measured 20 minutes after-lysis^42^ were associated with follow-up strains. Counterclockwise twist after-lysis was predictive of clockwise twisting at follow-up. Eyes with more glaucoma damage exhibited a more compliant strain response after-lysis and eyes with greater glaucoma damage at lysis experienced more axial expansion at follow-up. Together, these results suggest that the strain response and mechanical properties of the LC may provide biomarkers for the medium term remodeling process that could contribute to glaucoma damage. We hypothesize that strains measured over a short period of time (20 minutes after suturelysis or 1 week after IOP lowering by drops medication) could predict which patients will have greater glaucoma progressive worsening. Greater after-lysis shear strain had a borderline significant association with greater MD decrease at follow-up (p=0.12). It may be that a larger study will find that LC biomechanics offer a biomarker for the early diagnosis and progression of glaucoma.

A large follow-up *E_zz_* may indicate the LC is returning toward a state prior to damage, given that the LC is thinner and more curved with glaucoma damage.^18^ However, the eyes with the largest *E_zz_* and LC border movement had the thinnest RNFL thickness at the time of suturelysis (p=0.05, p=0.06 respectively). We previously reported that the short-term strain response (after-lysis) was more compliant in eyes with greater glaucoma damage, as measured by thinner RNF, more negative MD, and lower VFI.^42^ The result may be due to the thinner structure of a glaucoma LC, changes in the material properties of the LC with glaucoma damage, or a combination of both factors. While there has been some evidence that the sclera, overall, is less compliant with experimental glaucoma and in human glaucoma eyes,^46–53^ we determined that in vitro mechanical responses are more compliant in eyes with greater pre-mortem glaucoma damage.^54^ Furthermore, recent histological studies found that eyes with greater axon loss had thinner LC connective tissue beams.^19^ Taken together, these facts indicate that LC compliance is either greater in eyes destined to develop worse damage, or, that the damage process produces greater compliance. Prospective, longitudinal study of the predictive value of LC strains and their change over time are needed to determine which of these possibilities is more likely.

In our previous study, we defined LC compliance as strain per mm Hg IOP lowering, determining that greater compliance was associated with worse glaucoma damage.^42^ Since the IOP at follow-up was slightly, but not greatly lower than that immediately after suturelysis, we could not generate such a compliance measure for the eyes studied here. We did find that the IOP decrease at suturelysis was associated with greater follow-up LC border movement. However, there is a wide range of IOP change at suturelysis, which depends on a number of short-term factors in the immediate post-operative period. Generally, one might expect a larger mechanical stress (larger IOP change) to cause more movement in the short-term, but how this would influence more remote behavior is problematic.^20–24^

The main limitations of this study are its relatively small sample size, variable follow-up imaging times, and the variability and slight decrease in IOP at follow-up imaging. Initial patients in this study were imaged in 2018 and the repeat imaging was only undertaken after validation of the method.^34, 42^ Linear regression of the strain measures compared to time between did not show suturelysis and follow-up imaging trended toward a significant associations (p>0.06), and a larger study with multiple imaging time points is indicated. We plan to address both limitations by quantifying the changes in strain over time in eyes following eye drop treatment at consistent time intervals. In a subsequent report, we will demonstrate that valid strains can be measured one week after either starting or temporarily stopping eye drop treatment.

## Data Availability

All data produced in the present study are available upon reasonable request to the authors

## Abbreviations

LC: lamina cribrosa
ONH: optic nerve head
PPS: peripapillary sclera
SD-OCT: spectral domain optical coherence tomography
RNFL: retinal nerve fiber layer
MD: mean deviation
VFI: visual field index
IOP: intraocular pressure.

## Notes

**Conflict of Interest:** The authors have no conflicting relationships related to the devices used in this research. Heidelberg Engineering donated the imaging device, but were not involved with data analysis or interpretation, nor with drafting of the manuscript.

### Competing Interest Statement

The authors have no conflicting relationships related to the devices used in this research. Heidelberg Engineering donated the imaging device, but were not involved with data analysis or interpretation, nor with drafting of the manuscript.

### Clinical Trial

NCT03267849

### Author Declarations

The IRB of The Johns Hopkins Medicine gave ethical approval for this work.

