## Supplemental Tables for "Long-Term Remodeling Response in the Lamina Cribrosa Years after IOP Lowering by Suturelysis after Trabeculectomy"

### Supplemental Material

#### S1 DVC Errors

##### Supplemental Table S1: Average Baseline Error

| | Baseline $E_{rr}$ error | Baseline $E_{zz}$ error | Baseline $E_{rz}$ error | Baseline $E_{\theta\theta}$ error | Baseline $E_{r\theta}$ error | Baseline $E_{r\theta}$ error | Baseline $U_r$ error | Baseline $U_z$ error | Baseline $U_\theta$ error |
|---|---|---|---|---|---|---|---|---|---|
| LC072 | 0.0023 | 0.0023 | 0.0021 | 0.0019 | -0.00010 | -0.0046 | 0.44 | -0.90 | 0.63 |
| LC117 | -0.00043 | 0.0011 | -0.00019 | -0.000015 | -0.00046 | -0.0055 | -0.62 | -0.72 | -2.94 |
| LC167 | 0.0012 | 0.0027 | 0.0014 | -0.00037 | 0.0033 | -0.0053 | -0.72 | -0.70 | -3.79 |
| LC181 | 0.00091 | 0.0067 | 0.0028 | -0.0043 | -0.00048 | -0.0091 | -0.58 | -2.07 | 9.01 |
| LC182 | 0.00014 | 0.00083 | 0.00020 | 0.00083 | 0.000056 | 0.0056 | -0.12 | 0.20 | 3.51 |
| LC208 | -0.0012 | -0.00035 | 0.0012 | -0.0027 | 0.00025 | -0.0033 | 1.31 | -0.096 | 4.23 |
| LC229 | 0.0018 | 0.0010 | 0.00033 | 0.00082 | 0.0039 | -0.0096 | 0.81 | -0.018 | -0.28 |
| LC238 | 0.0000029 | 0.00022 | -0.00059 | -0.00018 | -0.00037 | 0.00016 | 0.26 | -0.075 | 0.17 |
| LC260 | -0.000067 | 0.00067 | -0.0020 | -0.0024 | -0.0012 | 0.000044 | 0.72 | -0.45 | 4.20 |
| LC296 | -0.00046 | 0.0017 | -0.00080 | -0.00067 | 0.00025 | -0.0052 | -0.59 | 0.082 | -0.065 |
| LC297 | 0.00044 | 0.0014 | -0.00049 | 0.0028 | 0.00049 | -0.0024 | -0.26 | 1.01 | 0.21 |
| LC322 | -0.00056 | 0.0016 | 0.0011 | 0.0016 | 0.00071 | -0.0024 | -0.22 | -1.20 | -1.26 |
| LC323 | -0.00084 | -0.0000070 | 0.0014 | 0.000091 | -0.0025 | 0.0014 | 0.65 | 0.021 | -0.35 |
| LC328 | -0.000042 | 0.00030 | 0.0012 | -0.0015 | 0.00027 | 0.0019 | 0.26 | 0.29 | -0.94 |
| LC356 | -0.0013 | 0.0013 | -0.00041 | -0.00046 | 0.00090 | 0.0042 | 0.48 | 0.20 | 0.97 |
| LC068 | 0.0015 | -0.000033 | -0.0013 | -0.0021 | -0.0017 | -0.0043 | 0.23 | 0.24 | 1.21 |

Supplemental Table S2: Average correlation error

| | Correlation $E_{rr}$ error | Correlation $E_{zz}$ error | Correlation $E_{rz}$ error | Correlation $E_{\theta\theta}$ error | Correlation $E_{r\theta}$ error | Correlation $E_{r\theta}$ error | Correlation $U_r$ error | Correlation $U_z$ error | Correlation $U_\theta$ error |
|---|---|---|---|---|---|---|---|---|---|
| LC072 | -0.0013 | 0.0040 | -0.00052 | 0.0028 | -0.000074 | -0.00095 | 0.012 | 0.74 | -0.35 |
| LC117 | -0.00067 | 0.0052 | 0.00017 | 0.0024 | 0.000082 | -0.00052 | -0.061 | 0.62 | 0.39 |
| LC167 | -0.0014 | 0.0037 | -0.000095 | 0.0028 | 0.00011 | -0.00028 | 0.052 | 0.65 | 0.27 |
| LC181 | -0.00071 | 0.0049 | -0.000019 | 0.0030 | 0.00043 | 0.00071 | -0.011 | 0.65 | -0.28 |
| LC182 | -0.00012 | 0.0055 | -0.00033 | 0.0035 | 0.000023 | -0.00030 | -0.0099 | 0.67 | 0.22 |
| LC208 | -0.0021 | 0.0091 | 0.00015 | 0.0040 | 0.00033 | 0.00015 | 0.068 | 0.57 | -0.067 |
| LC229 | -0.00067 | 0.0062 | 0.00024 | 0.0039 | 0.00013 | 0.0014 | -0.18 | 0.32 | -0.28 |
| LC238 | -0.00062 | 0.0043 | -0.000069 | 0.0031 | 0.000094 | 0.000019 | 0.051 | 0.63 | 0.14 |
| LC260 | -0.0012 | 0.0042 | -0.000082 | 0.0029 | 0.000068 | 0.00039 | -0.0037 | 0.73 | -0.12 |
| LC296 | -0.00046 | 0.0058 | 0.00052 | 0.0030 | 0.00012 | 0.00038 | -0.18 | 0.47 | -0.20 |
| LC297 | -0.00067 | 0.0052 | 0.000012 | 0.0028 | 0.0000066 | -0.00040 | -0.034 | 0.68 | 0.095 |
| LC322 | -0.0030 | 0.0051 | -0.0000029 | 0.0027 | -0.00060 | 0.0013 | -0.20 | 0.54 | 0.058 |
| LC323 | -0.00077 | 0.0055 | 0.000060 | 0.0026 | 0.0000079 | 0.00062 | 0.038 | 0.73 | -0.16 |
| LC328 | -0.0025 | 0.0054 | 0.00030 | 0.0026 | -0.000083 | -0.00018 | -0.070 | 0.61 | -0.15 |
| LC356 | -0.0012 | 0.0052 | -0.000056 | 0.0028 | 0.000097 | -0.0025 | 0.16 | 0.76 | 0.49 |
| LC068 | -0.0015 | 0.0055 | -0.000047 | 0.0023 | 0.0000011 | -0.00021 | -0.073 | 0.52 | -0.34 |

Supplemental Table S3: The specimen-averaged baseline and correlation errors (n = 16, 15 patients)

| | Strain Outcomes | | Baseline error | | | Correlation error | | |
|---|---|---|---|---|---|---|---|---|
| | Mean | std | mean | std | *p*-value | mean | std | *p*-value |
| $E_{zz}$ | 0.14 | 0.21 | 0.0013 | 0.0017 | **0.020** | 0.0053 | 0.0012 | **0.023** |
| $E_{rr}$ | 0.000099 | 0.014 | 0.00021 | 0.0011 | 0.97 | -0.0012 | 0.00079 | 0.72 |
| $E_{\theta\theta}$ | -0.0028 | 0.024 | -0.00042 | 0.0018 | 0.70 | 0.0029 | 0.00049 | 0.36 |
| $E_{r\theta}$ | 0.0048 | 0.013 | 0.00020 | 0.0016 | 0.17 | 0.000046 | 0.00022 | 0.16 |
| $E_{z\theta}$ | -0.0061 | 0.016 | -0.0024 | 0.0044 | 0.37 | -0.000023 | 0.00092 | 0.14 |
| $E_{rz}$ | 0.0050 | 0.026 | 0.00037 | 0.0013 | 0.50 | 0.000014 | 0.00024 | 0.45 |

**S2 LC depth change with strain**

Supplemental Table S4: Linear regression analysis comparing the follow-up anterior LC depth change to the follow-up strain. Also shown are *p*-values and associated $R^2$ value (n = 15, 14 patients).

| | Strain at no LC depth change | *p*-value | Change in strain per 1μm anterior LC depth change | *p*-value | $R^2$ |
|---|---|---|---|---|---|
| $E_{zz}$ | -0.032 | 0.58 | -0.0024 | **0.0044** | 0.48 |
| $E_{rr}$ | 0.0015 | 0.79 | 0.000031 | 0.67 | 0.015 |
| $E_{\theta\theta}$ | 0.0031 | 0.73 | 0.00014 | 0.23 | 0.11 |
| $E_{r\theta}$ | 0.0053 | 0.30 | 0.000031 | 0.62 | 0.019 |
| $E_{z\theta}$ | -0.011 | 0.056 | -0.00011 | 0.11 | 0.18 |
| $E_{rz}$ | 0.00024 | 0.98 | -0.000028 | 0.82 | 0.0042 |
| $E_{max}$ | 0.0099 | 0.85 | -0.0023 | **0.0028** | 0.51 |
| $\Gamma_{max}$ | 0.025 | 0.31 | -0.0011 | **0.0019** | 0.54 |

**S3 LC after-lysis and IOP change**

Supplemental Table S5: Linear regression analysis comparing the IOP decrease from before to 20 minutes after suturelysis to the follow-up strain. Also shown are *p*-values and associated $R^2$ value (Strain n = 16, 15 patients, LC Depth n = 15, 14 patients).

| | Strain at 0 IOP | *p*-value | Change in strain per 1mmHg IOP decrease | *p*-value | $R^2$ |
|---|---|---|---|---|---|
| *LC Depth* | -94.81 | 0.00035 | 2.50 | **0.046** | 0.27 |
| $E_{zz}$ | 0.23 | 0.018 | -0.0063 | 0.22 | 0.11 |
| $E_{rr}$ | -0.0018 | 0.76 | 0.00014 | 0.68 | 0.013 |
| $E_{\theta\theta}$ | -0.0035 | 0.74 | 0.000053 | 0.93 | 0.00057 |
| $E_{r\theta}$ | 0.0076 | 0.17 | -0.00021 | 0.51 | 0.031 |
| $E_{z\theta}$ | -0.0037 | 0.58 | -0.00018 | 0.64 | 0.016 |
| $E_{rz}$ | 0.012 | 0.30 | -0.00048 | 0.45 | 0.042 |
| $E_{max}$ | 0.25 | 0.0076 | -0.0055 | 0.26 | 0.090 |
| $\Gamma_{max}$ | 0.14 | 0.0030 | -0.0024 | 0.30 | 0.077 |

Supplemental Table S6: Linear regression analysis comparing the IOP decrease from before trabeculectomy to after-lysis to the follow-up strain. Also shown are *p*-values and associated $R^2$ value (Strain n = 16, 15 patients, LC Depth n = 15, 14 patients).

| | Strain at 0 IOP | *p*-value | Change in strain per 1mmHg IOP decrease | *p*-value | $R^2$ |
|---|---|---|---|---|---|
| *LC Depth* | -65.71 | 0.0029 | 0.691 | 0.66 | 0.016 |
| $E_{zz}$ | 0.16 | 0.040 | -0.0022 | 0.72 | 0.0095 |
| $E_{rr}$ | -0.0044 | 0.29 | 0.00064 | 0.084 | 0.20 |
| $E_{\theta\theta}$ | -0.011 | 0.13 | 0.0012 | 0.060 | 0.23 |
| $E_{r\theta}$ | 0.0021 | 0.61 | 0.00038 | 0.28 | 0.082 |
| $E_{z\theta}$ | -0.0092 | 0.080 | 0.00044 | 0.31 | 0.072 |
| $E_{rz}$ | 0.0068 | 0.43 | -0.00026 | 0.73 | 0.0088 |
| $E_{max}$ | 0.19 | 0.010 | -0.0022 | 0.70 | 0.011 |
| $\Gamma_{max}$ | 0.12 | 0.0021 | -0.0015 | 0.60 | 0.021 |

Supplemental Table S7: Linear regression analysis comparing the IOP decrease from after-lysis to follow-up to the follow-up strain. Also shown are *p*-values and associated $R^2$ value (Strain n = 16, 15 patients, LC Depth n = 15, 14 patients).

| | Strain at 0 IOP | *p*-value | Change in strain per 1mmHg IOP decrease | *p*-value | $R^2$ |
|---|---|---|---|---|---|
| *LC Depth* | -49.49 | 0.0054 | -2.85 | 0.11 | 0.18 |
| $E_{zz}$ | 0.092 | 0.13 | 0.012 | 0.096 | 0.19 |
| $E_{rr}$ | 0.0030 | 0.44 | -0.00069 | 0.13 | 0.16 |
| $E_{\theta\theta}$ | 0.00059 | 0.93 | -0.00082 | 0.32 | 0.070 |
| $E_{r\theta}$ | 0.0045 | 0.26 | 0.000078 | 0.86 | 0.0023 |
| $E_{z\theta}$ | -0.0052 | 0.27 | -0.00023 | 0.67 | 0.013 |
| $E_{rz}$ | 0.00010 | 0.99 | 0.0012 | 0.17 | 0.13 |
| $E_{max}$ | 0.13 | 0.028 | 0.011 | 0.11 | 0.17 |
| $\Gamma_{max}$ | 0.086 | 0.0052 | 0.0053 | 0.099 | 0.18 |

Supplemental Table S8: Linear regression analysis comparing the years between after-lysis and follow-up to the follow-up strain. Also shown are *p*-values and associated $R^2$ value (Strain n = 16, 15 patients, LC Depth n = 15, 14 patients).

| | Strain at 0 years | *p*-value | Change in strain per 1 year | *p*-value | $R^2$ |
|---|---|---|---|---|---|
| *LC Depth* | -103.07 | 0.012 | 22.13 | 0.21 | 0.12 |
| $E_{zz}$ | 0.094 | 0.48 | 0.023 | 0.70 | 0.011 |
| $E_{rr}$ | 0.0011 | 0.90 | -0.00050 | 0.90 | 0.0013 |
| $E_{\theta\theta}$ | -0.025 | 0.081 | 0.011 | 0.086 | 0.20 |
| $E_{r\theta}$ | -0.0033 | 0.67 | 0.0040 | 0.26 | 0.090 |
| $E_{z\theta}$ | 0.0094 | 0.28 | -0.0076 | 0.061 | 0.23 |
| $E_{rz}$ | -0.00040 | 0.98 | 0.0027 | 0.71 | 0.0099 |
| $E_{max}$ | 0.12 | 0.35 | 0.028 | 0.62 | 0.018 |
| $\Gamma_{max}$ | 0.073 | 0.24 | 0.017 | 0.53 | 0.028 |

Supplemental Table S9: Linear regression analysis comparing the mean IOP recorded between after-lysis to follow-up to the follow-up strain. Also shown are *p*-values and associated $R^2$ value (Strain n = 16, 15 patients, LC Depth n = 15, 14 patients).

| | Strain at 0 IOP | *p*-value | Change in strain per 1mmHg IOP | *p*-value | $R^2$ |
|---|---|---|---|---|---|
| *LC Depth* | -110.42 | 0.020 | 4.37 | 0.23 | 0.11 |
| $E_{zz}$ | 0.30 | 0.069 | -0.015 | 0.28 | 0.082 |
| $E_{rr}$ | -0.0016 | 0.88 | 0.00016 | 0.86 | 0.0022 |
| $E_{\theta\theta}$ | -0.015 | 0.42 | 0.0011 | 0.48 | 0.036 |
| $E_{r\theta}$ | -0.0051 | 0.59 | 0.00090 | 0.27 | 0.085 |
| $E_{z\theta}$ | -0.00037 | 0.97 | -0.00052 | 0.60 | 0.020 |
| $E_{rz}$ | 0.021 | 0.30 | -0.0014 | 0.40 | 0.051 |
| $E_{max}$ | 0.31 | 0.051 | -0.012 | 0.34 | 0.066 |
| $\Gamma_{max}$ | 0.16 | 0.037 | -0.0052 | 0.41 | 0.050 |

Supplemental Table S10: Linear regression analysis comparing the maximum IOP recorded between after-lysis to follow-up to the follow-up strain. Also shown are *p*-values and associated $R^2$ value (Strain n = 16, 15 patients, LC Depth n = 15, 14 patients).

| | Strain at 0 IOP | *p*-value | Change in strain per 1mmHg IOP | *p*-value | $R^2$ |
|---|---|---|---|---|---|
| *LC Depth* | -74.43 | 0.037 | 0.65 | 0.64 | 0.017 |
| $E_{zz}$ | 0.19 | 0.15 | -0.0022 | 0.68 | 0.012 |
| $E_{rr}$ | 0.00067 | 0.93 | -0.000028 | 0.94 | 0.00048 |
| $E_{\theta\theta}$ | -0.014 | 0.31 | 0.00056 | 0.36 | 0.060 |
| $E_{r\theta}$ | -0.0069 | 0.30 | 0.00058 | 0.060 | 0.23 |
| $E_{z\theta}$ | 0.0018 | 0.84 | -0.00039 | 0.32 | 0.071 |
| $E_{rz}$ | -0.0066 | 0.65 | 0.00057 | 0.38 | 0.055 |
| $E_{max}$ | 0.22 | 0.081 | -0.0019 | 0.71 | 0.0099 |
| $\Gamma_{max}$ | 0.12 | 0.043 | -0.00076 | 0.76 | 0.0070 |

Supplemental Table S11: Linear regression analysis comparing the minimum IOP recorded between after-lysis to follow-up to the follow-up strain. Also shown are *p*-values and associated $R^2$ value (Strain n = 16, 15 patients, LC Depth n = 15, 14 patients).

| | Strain at 0 IOP | *p*-value | Change in strain per 1mmHg IOP | *p*-value | $R^2$ |
|---|---|---|---|---|---|
| *LC Depth* | -86.95 | 0.0070 | 4.40 | 0.28 | 0.087 |
| $E_{zz}$ | 0.22 | 0.048 | -0.014 | 0.38 | 0.056 |
| $E_{rr}$ | -0.0037 | 0.58 | 0.00067 | 0.51 | 0.032 |
| $E_{\theta\theta}$ | -0.0075 | 0.54 | 0.00083 | 0.64 | 0.016 |
| $E_{r\theta}$ | 0.0016 | 0.79 | 0.00055 | 0.56 | 0.025 |
| $E_{z\theta}$ | -0.0070 | 0.37 | 0.00015 | 0.89 | 0.0013 |
| $E_{rz}$ | 0.020 | 0.11 | -0.0027 | 0.15 | 0.14 |
| $E_{max}$ | 0.25 | 0.021 | -0.013 | 0.39 | 0.053 |
| $\Gamma_{max}$ | 0.14 | 0.0082 | -0.0061 | 0.39 | 0.054 |

Supplemental Table S12: Linear regression analysis comparing the range of IOPs recorded between after-lysis to follow-up to the follow-up strain. Also shown are *p*-values and associated $R^2$ value (Strain n = 16, 15 patients, LC Depth n = 15, 14 patients).

| | Strain at 0 IOP | *p*-value | Change in strain per 1mmHg IOP | *p*-value | $R^2$ |
|---|---|---|---|---|---|
| *LC Depth* | -63.01 | 0.023 | 0.14 | 0.91 | 0.00092 |
| $E_{zz}$ | 0.15 | 0.13 | -0.00056 | 0.91 | 0.00085 |
| $E_{rr}$ | 0.0015 | 0.80 | -0.000099 | 0.77 | 0.0064 |
| $E_{\theta\theta}$ | -0.0090 | 0.40 | 0.00043 | 0.47 | 0.038 |
| $E_{r\theta}$ | -0.0021 | 0.68 | 0.00047 | 0.12 | 0.17 |
| $E_{z\theta}$ | -0.00061 | 0.93 | -0.00038 | 0.31 | 0.072 |
| $E_{rz}$ | -0.0070 | 0.52 | 0.00082 | 0.18 | 0.12 |
| $E_{max}$ | 0.18 | 0.058 | -0.00036 | 0.94 | 0.00040 |
| $\Gamma_{max}$ | 0.11 | 0.024 | -0.000035 | 0.99 | 0.000016 |

Supplemental Table S13: Linear regression analysis comparing the standard deviation of IOPs recorded between after-lysis to follow-up to the follow-up strain. Also shown are *p*-values and associated $R^2$ value (Strain n = 16, 15 patients, LC Depth n = 15, 14 patients).

| | Strain at 0 IOP | *p*-value | Change in strain per 1mmHg IOP | *p*-value | $R^2$ |
|---|---|---|---|---|---|
| *LC Depth* | -68.20 | 0.020 | 1.55 | 0.74 | 0.0090 |
| $E_{zz}$ | 0.18 | 0.092 | -0.0077 | 0.67 | 0.014 |
| $E_{rr}$ | 0.0013 | 0.83 | -0.00027 | 0.81 | 0.0041 |
| $E_{\theta\theta}$ | -0.0072 | 0.53 | 0.00096 | 0.64 | 0.017 |
| $E_{r\theta}$ | -0.00040 | 0.94 | 0.0011 | 0.28 | 0.081 |
| $E_{z\theta}$ | -0.00069 | 0.92 | -0.0012 | 0.36 | 0.061 |
| $E_{rz}$ | -0.0057 | 0.63 | 0.0023 | 0.28 | 0.084 |
| $E_{max}$ | 0.21 | 0.038 | -0.0073 | 0.67 | 0.014 |
| $\Gamma_{max}$ | 0.12 | 0.015 | -0.0033 | 0.68 | 0.012 |

Supplemental Table S14: Linear regression analysis comparing the variance of IOPs recorded between after-lysis to follow-up to the follow-up strain. Also shown are *p*-values and associated $R^2$ value (Strain n = 16, 15 patients, LC Depth n = 15, 14 patients).

| | Strain at 0 IOP | *p*-value | Change in strain per 1mmHg IOP | *p*-value | $R^2$ |
|---|---|---|---|---|---|
| *LC Depth* | -61.76 | 0.0051 | 0.031 | 0.94 | 0.00049 |
| $E_{zz}$ | 0.14 | 0.053 | -0.00016 | 0.91 | 0.00089 |
| $E_{rr}$ | 0.0010 | 0.82 | -0.000034 | 0.73 | 0.0088 |
| $E_{\theta\theta}$ | -0.0054 | 0.50 | 0.000094 | 0.58 | 0.022 |
| $E_{r\theta}$ | 0.0013 | 0.74 | 0.00013 | 0.15 | 0.14 |
| $E_{z\theta}$ | -0.0040 | 0.43 | -0.000076 | 0.49 | 0.035 |
| $E_{rz}$ | -0.00069 | 0.93 | 0.00021 | 0.25 | 0.093 |
| $E_{max}$ | 0.18 | 0.014 | -0.00018 | 0.90 | 0.0012 |
| $\Gamma_{max}$ | 0.11 | 0.0036 | -0.000082 | 0.91 | 0.0010 |

**S4 LC after-lysis and glaucomatous neuropathy**

Supplemental Table S15: Linear regression analysis comparing the RNFL thickness measured at suturelysis to the follow-up strain. Also shown are *p*-values and associated $R^2$ value (Strain n = 16, 15 patients, LC Depth n = 15, 14 patients).

| | Strain at 0µm RNFL | *p*-value | Change in strain per 1µm RNFL | *p*-value | $R^2$ |
|---|---|---|---|---|---|
| *LC Depth* | -234.78 | 0.018 | 2.77 | 0.063 | 0.24 |
| $E_{zz}$ | 0.84 | 0.026 | -0.011 | **0.054** | 0.24 |
| $E_{rr}$ | 0.017 | 0.50 | -0.00027 | 0.50 | 0.033 |
| $E_{\theta\theta}$ | 0.0015 | 0.97 | -0.000068 | 0.92 | 0.00067 |
| $E_{r\theta}$ | 0.012 | 0.62 | -0.00011 | 0.77 | 0.0063 |
| $E_{z\theta}$ | 0.019 | 0.49 | -0.00041 | 0.36 | 0.060 |
| $E_{rz}$ | 0.064 | 0.17 | -0.00095 | 0.20 | 0.12 |
| $E_{max}$ | 0.81 | 0.026 | -0.010 | 0.070 | 0.22 |
| $\Gamma_{max}$ | 0.38 | 0.032 | -0.0044 | 0.11 | 0.17 |

Supplemental Table S16: Linear regression analysis comparing the MD measured at suturelysis to the follow-up strain. Also shown are *p*-values and associated $R^2$ value (Strain n = 16, 15 patients, LC Depth n = 15, 14 patients).

| | Strain at 0dB MD | *p*-value | Change in strain per 1dB MD | *p*-value | $R^2$ |
|---|---|---|---|---|---|
| *LC Depth* | -82.44 | 0.011 | -2.00 | 0.38 | 0.059 |
| $E_{zz}$ | 0.19 | 0.080 | 0.0049 | 0.56 | 0.024 |
| $E_{rr}$ | -0.0014 | 0.84 | -0.00014 | 0.80 | 0.0050 |
| $E_{\theta\theta}$ | -0.00086 | 0.94 | 0.00019 | 0.85 | 0.0027 |
| $E_{r\theta}$ | 0.0032 | 0.61 | -0.00015 | 0.77 | 0.0065 |
| $E_{z\theta}$ | -0.0061 | 0.42 | 0.000000033 | 1.00 | <0.0001 |
| $E_{rz}$ | 0.010 | 0.43 | 0.00050 | 0.63 | 0.017 |
| $E_{max}$ | 0.24 | 0.024 | 0.0062 | 0.44 | 0.043 |
| $\Gamma_{max}$ | 0.15 | 0.0057 | 0.0038 | 0.32 | 0.071 |

Supplemental Table S17: Linear regression analysis comparing the VFI measured at suturelysis to the follow-up strain. Also shown are *p*-values and associated $R^2$ value (Strain n = 16, 15 patients, LC Depth n = 15, 14 patients).

| | Strain at 0% VFI | *p*-value | Change in strain per 1% VFI | *p*-value | $R^2$ |
|---|---|---|---|---|---|
| *LC Depth* | -13.52 | 0.80 | -0.67 | 0.31 | 0.078 |
| $E_{zz}$ | 0.021 | 0.91 | 0.0016 | 0.52 | 0.031 |
| $E_{rr}$ | 0.0028 | 0.82 | -0.000038 | 0.82 | 0.0039 |
| $E_{\theta\theta}$ | -0.0013 | 0.95 | -0.000020 | 0.94 | 0.00036 |
| $E_{r\theta}$ | 0.0063 | 0.59 | -0.000021 | 0.89 | 0.0014 |
| $E_{z\theta}$ | -0.0056 | 0.69 | -0.0000070 | 0.97 | 0.00010 |
| $E_{rz}$ | -0.0046 | 0.85 | 0.00013 | 0.67 | 0.013 |
| $E_{max}$ | 0.034 | 0.85 | 0.0020 | 0.41 | 0.049 |
| $\Gamma_{max}$ | 0.022 | 0.79 | 0.0012 | 0.30 | 0.075 |

Supplemental Table S18: Linear regression analysis comparing the MD change from suturelysis to follow-up to the follow-up strain. Also shown are *p*-values and associated $R^2$ value (Strain n = 16, 15 patients, LC Depth n = 15, 14 patients).

| | Strain at 0dB MD | *p*-value | Change in strain per 1dB MD | *p*-value | $R^2$ |
|---|---|---|---|---|---|
| *LC Depth* | -61.07 | 0.0014 | -0.19 | 0.96 | 0.00021 |
| $E_{zz}$ | 0.14 | 0.033 | -0.0041 | 0.78 | 0.0060 |
| $E_{rr}$ | -0.00045 | 0.90 | -0.00049 | 0.60 | 0.020 |
| $E_{\theta\theta}$ | -0.0066 | 0.25 | -0.0034 | **0.024** | 0.31 |
| $E_{r\theta}$ | 0.0029 | 0.34 | -0.0017 | **0.033** | 0.28 |
| $E_{z\theta}$ | -0.0060 | 0.17 | 0.000078 | 0.94 | 0.00040 |
| $E_{rz}$ | 0.0018 | 0.78 | -0.0029 | 0.084 | 0.20 |
| $E_{max}$ | 0.17 | 0.0065 | -0.0031 | 0.82 | 0.0038 |
| $\Gamma_{max}$ | 0.11 | 0.0012 | -0.00082 | 0.90 | 0.0011 |

Supplemental Table S19: Linear regression analysis comparing the VFI change from suturelysis to follow-up to the follow-up strain. Also shown are *p*-values and associated $R^2$ value (Strain n = 16, 15 patients, LC Depth n = 15, 14 patients).

| | Strain at 0% VFI | *p*-value | Change in strain per 1% VFI | *p*-value | $R^2$ |
|---|---|---|---|---|---|
| *LC Depth* | -61.11 | 0.0014 | -0.0698 | 0.95 | 0.00029 |
| $E_{zz}$ | 0.14 | 0.027 | 0.00036 | 0.94 | 0.00046 |
| $E_{rr}$ | -0.00077 | 0.83 | -0.00026 | 0.37 | 0.058 |
| $E_{\theta\theta}$ | -0.0064 | 0.25 | -0.0011 | **0.020** | 0.33 |
| $E_{r\theta}$ | 0.0033 | 0.30 | -0.00044 | 0.088 | 0.19 |
| $E_{z\theta}$ | -0.0064 | 0.14 | -0.000094 | 0.77 | 0.0060 |
| $E_{rz}$ | 0.0037 | 0.60 | -0.00039 | 0.48 | 0.037 |
| $E_{max}$ | 0.18 | 0.0052 | 0.00061 | 0.89 | 0.0015 |
| $\Gamma_{max}$ | 0.11 | 0.00090 | 0.00057 | 0.78 | 0.0056 |

**S5 Comparison of after-lysis and follow-up strains**

Supplemental Table S20: Linear regression analysis comparing the after-lysis strain to the follow-up strain. Also shown are *p*-values and associated $R^2$ value (Strain n = 16, 15 patients).

| | Strain at 0 after-lysis strain | *p*-value | Change in follow-up strain per 1 after-lysis strain | *p*-value | $R^2$ |
|---|---|---|---|---|---|
| $E_{zz}$ | 0.15 | 0.037 | -1.42 | 0.71 | 0.010 |
| $E_{rr}$ | -0.0005 | 0.90 | -0.33 | 0.75 | 0.0073 |
| $E_{\theta\theta}$ | -0.0023 | 0.72 | 0.33 | 0.53 | 0.028 |
| $E_{r\theta}$ | 0.0045 | 0.21 | 0.13 | 0.79 | 0.0049 |
| $E_{z\theta}$ | -0.0074 | 0.032 | -1.67 | **0.0068** | 0.42 |
| $E_{rz}$ | 0.0050 | 0.47 | 0.21 | 0.87 | 0.0021 |
| $E_{max}$ | 0.21 | 0.017 | -1.62 | 0.57 | 0.024 |
| $\Gamma_{max}$ | 0.13 | 0.0046 | -1.48 | 0.42 | 0.046 |

Supplemental Table S21: Linear regression analysis comparing the strain after-lysis to the MD change from suturelysis to follow-up. Also shown are *p*-values and associated $R^2$ value (Strain n = 16, 15 patients, LC Depth n = 15, 14 patients).

| | MD at 0 strain | *p*-value | Change in MD per 1 after-lysis strain | *p*-value | $R^2$ |
|---|---|---|---|---|---|
| *LC Depth* | -1.20 | 0.24 | 0.18 | 0.21 | 0.11 |
| $E_{zz}$ | -1.49 | 0.25 | 36.18 | 0.61 | 0.019 |
| $E_{rr}$ | -0.63 | 0.59 | 260.70 | 0.39 | 0.053 |
| $E_{\theta\theta}$ | -1.20 | 0.26 | -54.38 | 0.53 | 0.029 |
| $E_{r\theta}$ | -0.65 | 0.52 | -238.72 | 0.12 | 0.16 |
| $E_{z\theta}$ | -1.05 | 0.32 | 75.58 | 0.67 | 0.013 |
| $E_{rz}$ | -1.11 | 0.30 | 17.57 | 0.93 | 0.00060 |
| $E_{max}$ | -1.09 | 0.50 | -1.21 | 0.98 | 0.000035 |
| $\Gamma_{max}$ | -0.58 | 0.73 | -31.19 | 0.68 | 0.012 |
